# Trends in SARS-CoV-2 seroprevalence in Massachusetts estimated from newborn screening specimens

**DOI:** 10.1101/2021.10.29.21265678

**Authors:** Kevin C. Ma, Jaime E. Hale, Yonatan H. Grad, Galit Alter, Katherine Luzuriaga, Roger B. Eaton, Stephanie Fischinger, Devinder Kaur, Robin Brody, Sameed M. Siddiqui, Dylan Leach, Catherine M. Brown, R. Monina Klevens, Lawrence Madoff, Anne Marie Comeau

## Abstract

**Background:** Estimating the cumulative incidence of SARS-CoV-2 is essential for setting public health policies. We leveraged de-identified Massachusetts newborn screening specimens to generate an accessible, retrospective source of maternal antibodies for estimating statewide SARS-CoV-2 seroprevalence in a non-test-seeking population.

**Methods:** We analyzed 72,117 newborn dried blood spots collected from November 2019 through December 2020, representing 337 towns and cities across Massachusetts. Seroprevalence was estimated for the general Massachusetts population after correcting for imperfect test specificity and nonrepresentative sampling using Bayesian multilevel regression and poststratification.

**Results:** Statewide seroprevalence was estimated to be 0.03% (90% credible interval (CI) [0.00, 0.11]) in November 2019 and rose to 1.47% (90% CI [1.00, 2.13]) by May 2020, following sustained SARS-CoV-2 transmission in the spring. Seroprevalence plateaued from May onwards, reaching 2.15% (90% CI [1.56, 2.98]) in December 2020. Seroprevalence varied substantially by community and was particularly associated with community percent non-Hispanic Black (β = 0.024, 90% CI [0.004, 0.044]); i.e., a 10% increase in community percent non-Hispanic Black was associated with a 27% higher odds of seropositivity. Seroprevalence estimates had good concordance with reported case counts and wastewater surveillance for most of 2020, prior to the resurgence of transmission in winter.

**Conclusions:** Cumulative incidence of SARS-CoV-2 protective antibody in Massachusetts was low as of December 2020, indicating that a substantial fraction of the population was still susceptible. Maternal seroprevalence data from newborn screening can inform longitudinal trends and identify cities and towns at highest risk, particularly in settings where widespread diagnostic testing is unavailable.

**Summary:** The measurement of maternal antibodies in dried blood spots collected for newborn screening offers a statewide source of SARS-CoV-2 seroprevalence data independent of case testing limitations. We analyzed 72,117 Massachusetts spots collected November 2019 – December 2020 and estimated longitudinal trends.

## Introduction

Disease surveillance networks operate at multiple levels of government and healthcare, yet monitoring for outbreaks of new diseases remains a challenge. There are few material sources of information with limited cohort bias available to address questions about the frequency and distribution of a new infectious agent within populations. Newborn screening (NBS) programs are one of those resources. A successful public health service, NBS programs collect infant dried blood spot specimens at centralized clinical laboratories where they are tested for markers of an ever-expanding panel of treatable, largely genetic, disorders and referred to appropriate providers when necessary. Such programs comprise high-throughput laboratory testing with community outreach, operate under state authority, and maintain secure electronic and specimen records. Importantly, these infant blood specimens collected for NBS contain maternal IgG antibodies that cross the placenta and that reflect maternal exposure to infectious agents. The use of NBS to measure the seroprevalence of an emerging infectious disease was pioneered in the late 1980s for HIV [1] and taken up by a majority of states for several years [2].

This framework offers advantages for monitoring SARS-CoV-2 infection in the absence of symptomatic disease, particularly as diagnostic testing was limited in the initial stages of the pandemic, leading to uncertainty about the true burden and spread of infection. Many critical questions about SARS-CoV-2 infection incidence and spread are important to answer to inform public health responses to the pandemic: What proportion of our population has been exposed? How is exposure geographically distributed? Are the rates of exposure increasing in some populations faster than in others? What are the characteristics of those who become infected? Using data generated from NBS specimens, we report findings that address these questions from a retrospective, de-identified, and systematic survey of SARS-CoV-2 seroprevalence in childbearing women in Massachusetts.

## Methods

### Study population

Women who were residents of Massachusetts, gave birth in Massachusetts and whose infants’ dried blood spot (DBS) specimens had completed routine newborn screening met study surveillance inclusion criteria with the DBS specimens serving as surrogates for the women. A control group was pre-defined to be those DBS arriving at the newborn screening program in March of 2019 and the study group was defined as those DBS arriving from November 4, 2019 through December 31, 2020. The following exclusion criteria were applied: only one DBS specimen per infant and only one DBS specimen from each multiples’ birth was included. All specimens of infants >30 days of age at time of specimen, all DBS of infants transfused within 48 hours, all specimens determined to be “unsatisfactory” quality for NBS, and all specimens from mothers who opted out of providing data for research were excluded.

The Institutional Review Boards of the Massachusetts Department of Public Health and the UMass Chan Medical School approved waivers of consent for the de-identified public health surveillance.

### Antibody testing optimization

Human monoclonal IgG antibody cross reactive to SARS-CoV-2 was prepared from CR3022 variable genes expressed from plasmids, as described previously for other SARS-CoV-2 antibodies [3]. The receptor binding domain (RBD) of the Spike protein used as ELISA Ag was expressed using HEK-293F cells and prepared as follows: a plasmid containing His-tagged RBD was transiently transfected in HEK-293F cells using PEI. The expressed protein was extracted and purified with Ni-NTA resin and stored at -80°C until use. IgG antibodies specific to the RBD protein were detected in eluates from DBSs residual to the Massachusetts newborn screening program, by an adaptation of a previously described ELISA designed for serum [4]. Details of the protocol used can be found in the Supplementary Methods.

### Cutoff determination

To determine positive and negative interpretations, we used a plate-specific cutoff calculation using standard deviations (SD) above the mean of the replicates’ average IgG concentration in a two-step process. In the first step, the mean and two SD were calculated for each plate. In the second step, any specimens with a concentration greater than or equal to two standard deviations above the plate mean from the first step were excluded from the calculation of the second mean and standard deviation (mean and SD of presumed negative samples). For Laboratory 1 (384-well assays), any specimen with a µg/mL result that was 5.3 SD above the second cycle mean was interpreted as positive. For laboratories 2 and 3 (96 well assays), any specimen with a µg/mL result that was 3.3 SD above the second cycle mean was interpreted as positive.

### Inhibition assay

The inhibition assay was performed in the same manner as the standard IgG antibody analysis, with the following exceptions. Concentrated RBD diluted in dilution buffer was pre-mixed with DBS eluates so that the final dilution of the DBS eluates was identical to that used in the standard assay (1:4), and the final dilution of RBD was 1 µg/ml. This mixture was incubated for 30 minutes at RT, and 100 µl of the inhibited mixture was added to ELISA wells in duplicate. Uninhibited wells were also tested in duplicate, replacing the concentrated RBD with diluent alone. Percent inhibition was expressed as 100 * (average OD of inhibited wells) / (average OD of uninhibited wells).

### Data processing

Each of three testing laboratories determined that test results met Quality Control (QC) requirements. Testing laboratories reported replicate OD and IgG concentration results for all well locations from specified plates to investigators at the New England Newborn Screening Program (NENSP). Transient linkages between plated specimens, test results, and demographic data were created and available to only two investigators at the NENSP (AMC and JEH) for application of inclusion and exclusion criteria followed by calculations for results interpretation in the restricted study database. Data extracted by a query of the restricted database yielded a de-identified set of coded specimens with the limited data variables reported in this paper. All transient linkages to any identifiers will be irretrievably destroyed upon acceptance for publication.

### Population-wide statistical model

We used dynamic (i.e., time-varying) Bayesian multilevel regression and poststratification (MRP) models to adjust seroprevalence estimates for non-representative sampling by age, sex, and Massachusetts community of residence (i.e., n=351 cities or towns) and for imperfect test specificity [5-9]. The core approach of MRP is to estimate the outcome variable conditional on sociodemographic characteristics using a non-representative data set, and then project these estimates onto a representative population of interest [5, 7-11]. MRP models additionally enable estimation of longitudinal seroprevalence trends at granular geographic levels, such as the communities that we consider here. Estimating these models consists of two steps: first, we fit a time-varying Bayesian multilevel logistic regression model to predict serostatus conditional on age category and community; these covariates are modeled using random effects [10]. We used two types of time-varying functions: we primarily assessed estimates from a model grouping data by month, but for comparison, we also fit a generalized additive model using thin-plate splines which modeled time continuously [8]. Then, we post-stratified the seroprevalence estimates over time using census data to individual communities and to the statewide population. In the monthly model, we also accounted for imperfect test specificity by allowing for measurement error when collecting data from true seronegatives [9]. Additional details for the models used can be found in the Supplementary Methods.

To identify community-level factors associated with seropositivity, we also fit a time-invariant logistic regression model pooling data from the last four months (i.e., September through December of 2020, corresponding to the height of seropositivity). We included fixed effects for multiple demographic factors acquired from the 2015-2019 5-year American Census Survey (Supplementary Table 2) [12] and random effects for age category and community. Details for all model fitting parameters can be found in the Supplementary Methods. Code used to fit the models will be made available on Github at the following URL: https://github.com/gradlab/covid19-newborn-seroprevalence

### Other data sources

We acquired deidentified active case surveillance incidence data from the Massachusetts Department of Public Health’s MAVEN, an integrated, web-based disease surveillance and case management system [13]. We also acquired data from wastewater surveillance generated by the Massachusetts Water Resource Authority (MWRA) and Biobot Analytics [14]. Because seroprevalence estimates are a proxy for cumulative incidence up until a given time, we took the cumulative sum of the MAVEN incidence or MWRA wastewater surveillance data, respectively. To account for the delay from exposure to SARS-CoV-2 to antibody generation, we lagged the seroprevalence model by 3 weeks when overlaying longitudinal trends [15].

## Results

Neonatal specimens representing all Massachusetts women who gave birth and met study criteria are shown in Table 1. The 72,117 women who gave birth from November 2019 through December 2020 resided in 337 towns and cities across Massachusetts. The 1,817 presumed seronegative (March 2019) specimens were also from across the state (324 towns and cities).

**Table 1.**
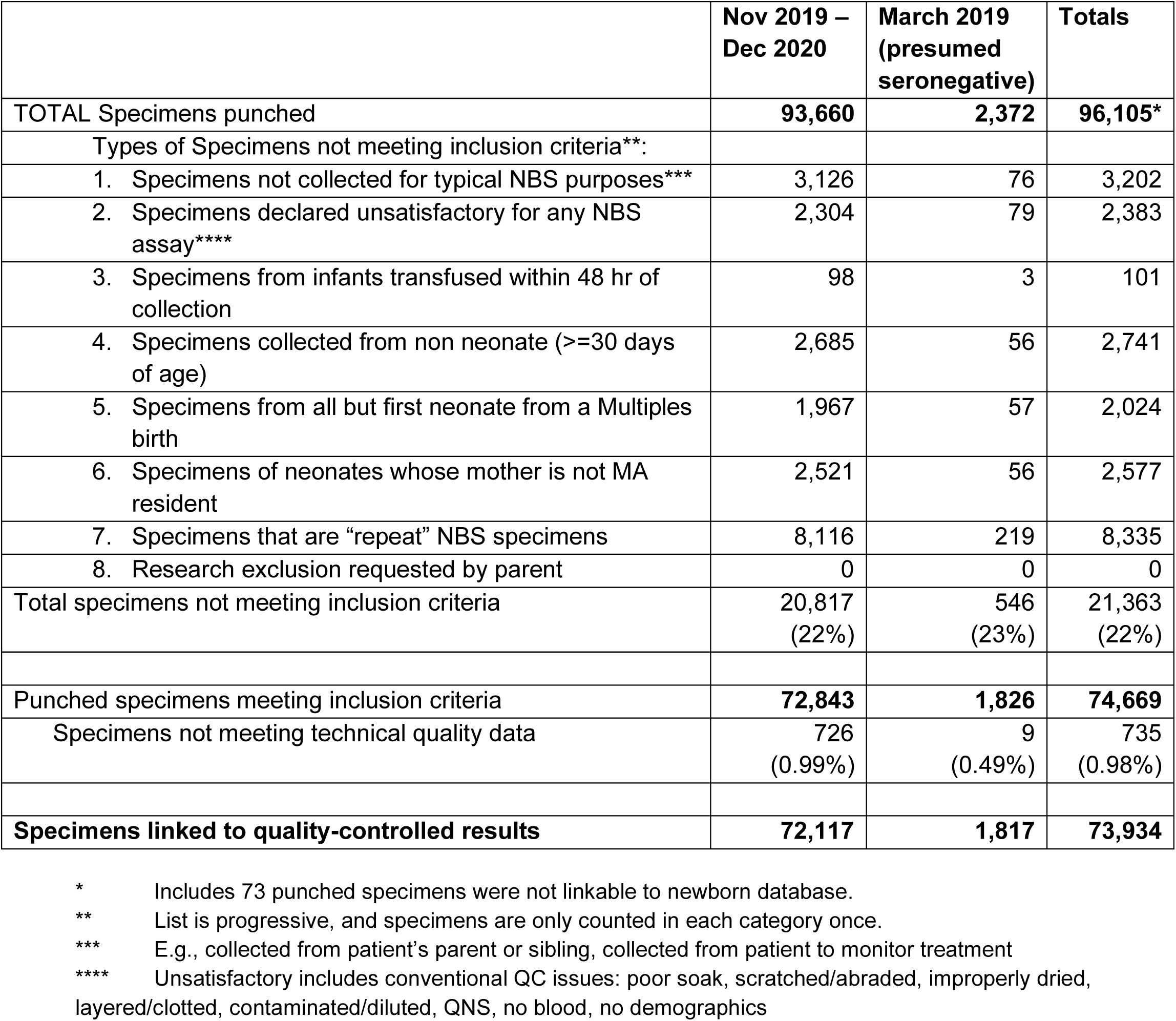
Characteristics of NBS DBS specimens included in the survey.

|  | Nov 2019 –<br>Dec 2020 | March 2019<br>(presumed<br>seronegative) | Totals |
| --- | --- | --- | --- |
| TOTAL Specimens punched | **93,660** | **2,372** | **96,105\*** |
| Types of Specimens not meeting inclusion criteria**: |  |  |  |
| 1. Specimens not collected for typical NBS purposes*** | 3,126 | 76 | 3,202 |
| 2. Specimens declared unsatisfactory for any NBS assay**** | 2,304 | 79 | 2,383 |
| 3. Specimens from infants transfused within 48 hr of collection | 98 | 3 | 101 |
| 4. Specimens collected from non neonate ($\geq 30$ days of age) | 2,685 | 56 | 2,741 |
| 5. Specimens from all but first neonate from a Multiples birth | 1,967 | 57 | 2,024 |
| 6. Specimens of neonates whose mother is not MA resident | 2,521 | 56 | 2,577 |
| 7. Specimens that are “repeat” NBS specimens | 8,116 | 219 | 8,335 |
| 8. Research exclusion requested by parent | 0 | 0 | 0 |
| Total specimens not meeting inclusion criteria | 20,817<br>(22%) | 546<br>(23%) | 21,363<br>(22%) |
| Punched specimens meeting inclusion criteria | **72,843** | **1,826** | **74,669** |
| Specimens not meeting technical quality data | 726<br>(0.99%) | 9<br>(0.49%) | 735<br>(0.98%) |
| **Specimens linked to quality-controlled results** | **72,117** | **1,817** | **73,934** |

\* Includes 73 punched specimens were not linkable to newborn database.
\*\* List is progressive, and specimens are only counted in each category once.
\*\*\* E.g., collected from patient’s parent or sibling, collected from patient to monitor treatment
\*\*\*\* Unsatisfactory includes conventional QC issues: poor soak, scratched/abraded, improperly dried, layered/clotted, contaminated/diluted, QNS, no blood, no demographics

**Table 2.**
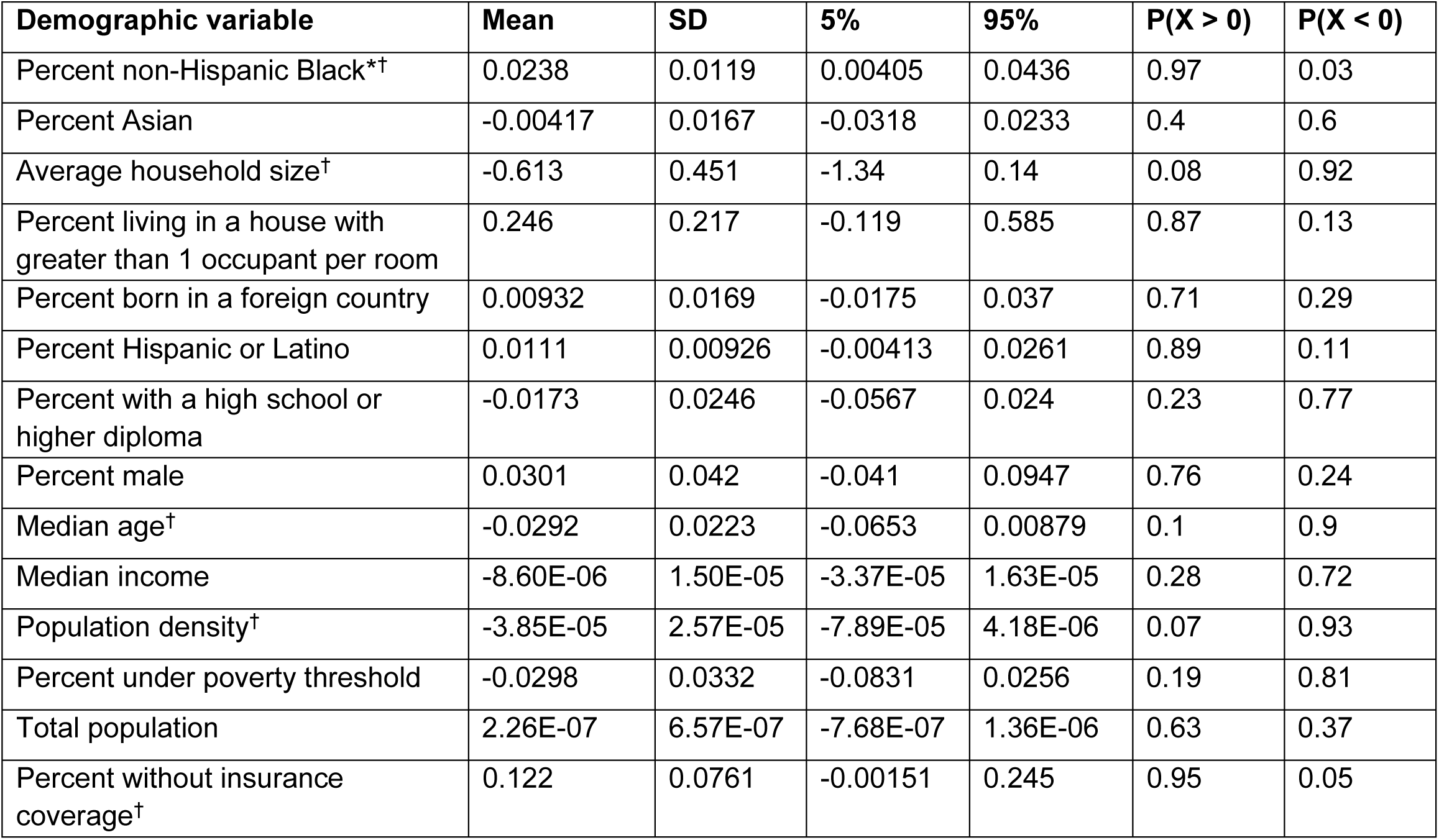
Posterior distribution summaries for the community-level multivariate association of demographic characteristics with seroprevalence. † indicates either P(X > 0) or P(X < 0) is 90% or higher and * indicates that the 90% credible interval excludes 0.

We first measured seropositive rates from a population of anonymized newborn DBS in a subset of residual DBS from a period believed to significantly pre-date the COVID-19 outbreak (March 2019). Each of the three testing labs received punches from the same 288 specimens punched across 4 plates. Figure 1 shows the individual [IgG] results of the anonymized presumed seronegative specimens, the positive controls, and diluent controls by plate and by testing laboratory. The low positive controls showed IgG concentrations significantly above the mean of the tested specimens and all above the values for the tested specimens.

**Figure 1.**
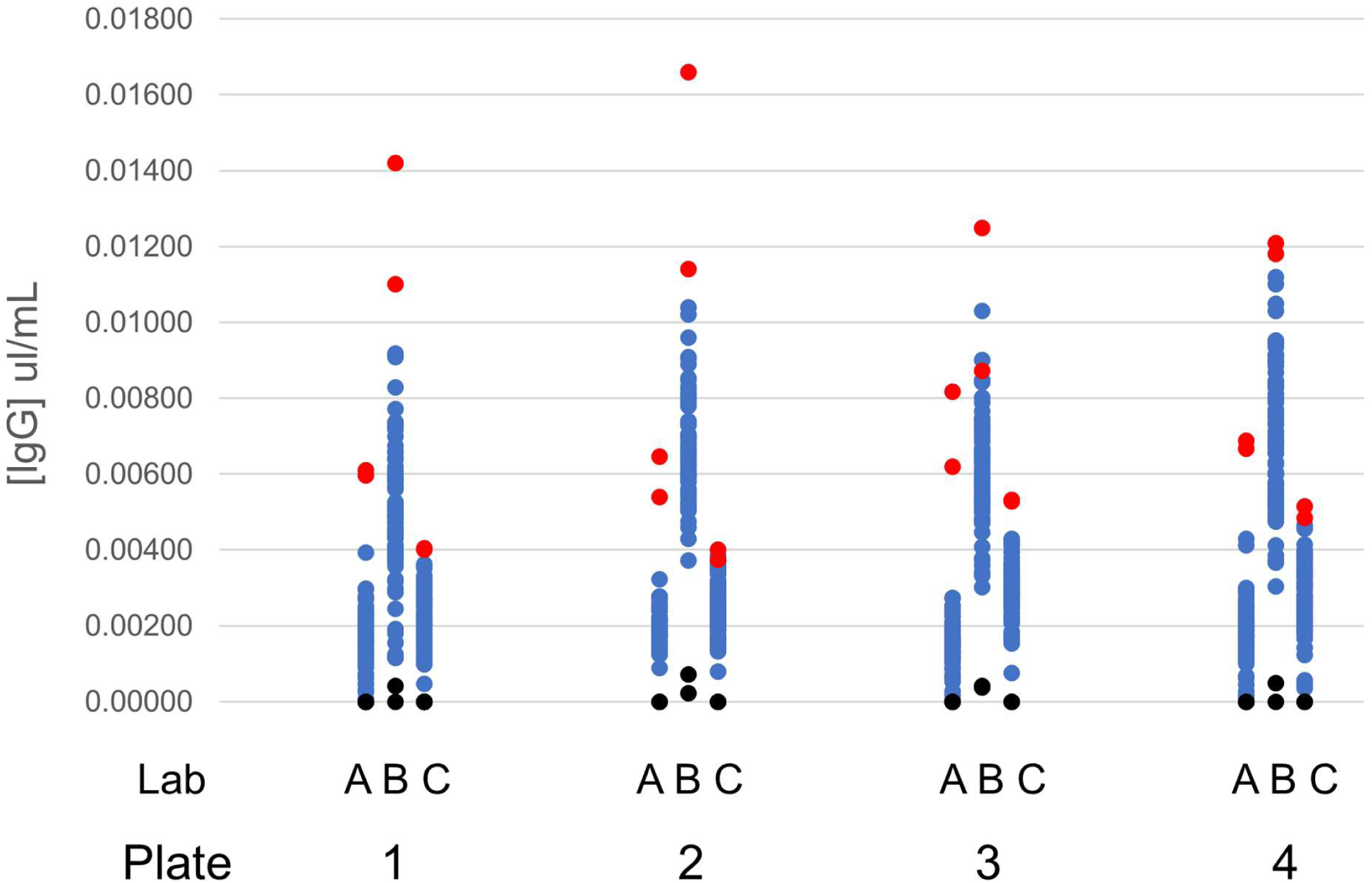
[IgG] results of 288 anonymized specimens obtained in March 2019 tested by each of the three labs across four plates. Blue dots are residual NBS DBS specimens, red dots are positive controls, and black dots are diluent controls.

Among the 1,817 presumed seronegative specimens from March 2019, 7 (0.39%) were seropositive for SARS-CoV-2. Among the 72,117 study specimens, 1,261 (1.75% statewide) were seropositive; 45 seropositives were from November and December 2019. We investigated the likelihood that these early seropositive values were false positives using an inhibition assay. For comparison, of the 29 seropositive specimens from July 2020, the average inhibition was 57% (confidence interval [48, 66]; minimum 22%). Of the available 45 specimens from November and December 2019, the average inhibition was 1.45% (confidence interval [1.34, 1.56]), confirming that most seropositives from 2019 were false positives. Only 3 of the 45 specimens showed notable inhibition (18%, 27%, and 36%).

We projected the NBS seroprevalence results statewide and to Massachusetts cities and towns using multilevel regression and poststratification (MRP) and additionally accounted for imperfect test specificity. MRP enables estimation of seroprevalence at granular geographic levels and addresses non-representative sampling, which is a challenge for the NBS cohort since there are sampling biases by sex, age group, and geography relative to the overall Massachusetts population (Supplementary Figures 1 and 2). Statewide monthly seroprevalence in early November 2019 was estimated to be 0.03% with a 90% credible interval (CI) of 0.00% to 0.11% and remained low until May 2020 (Figure 2). In May 2020, seroprevalence rose to 1.47% (90% CI [1.00, 2.13]) following sustained SARS-CoV-2 transmission in the spring, and plateaued at approximately 2% from July onwards, reflecting decreased transmission in the summer months due to lockdowns, mask wearing, and other factors [16, 17]. The estimate for seroprevalence for December 2020 was 2.15% (90% CI [1.56, 2.98]). Statewide trends estimated from the continuous-time MRP model showed similar qualitative results (Supplementary Figure 3).

**Figure 2.**
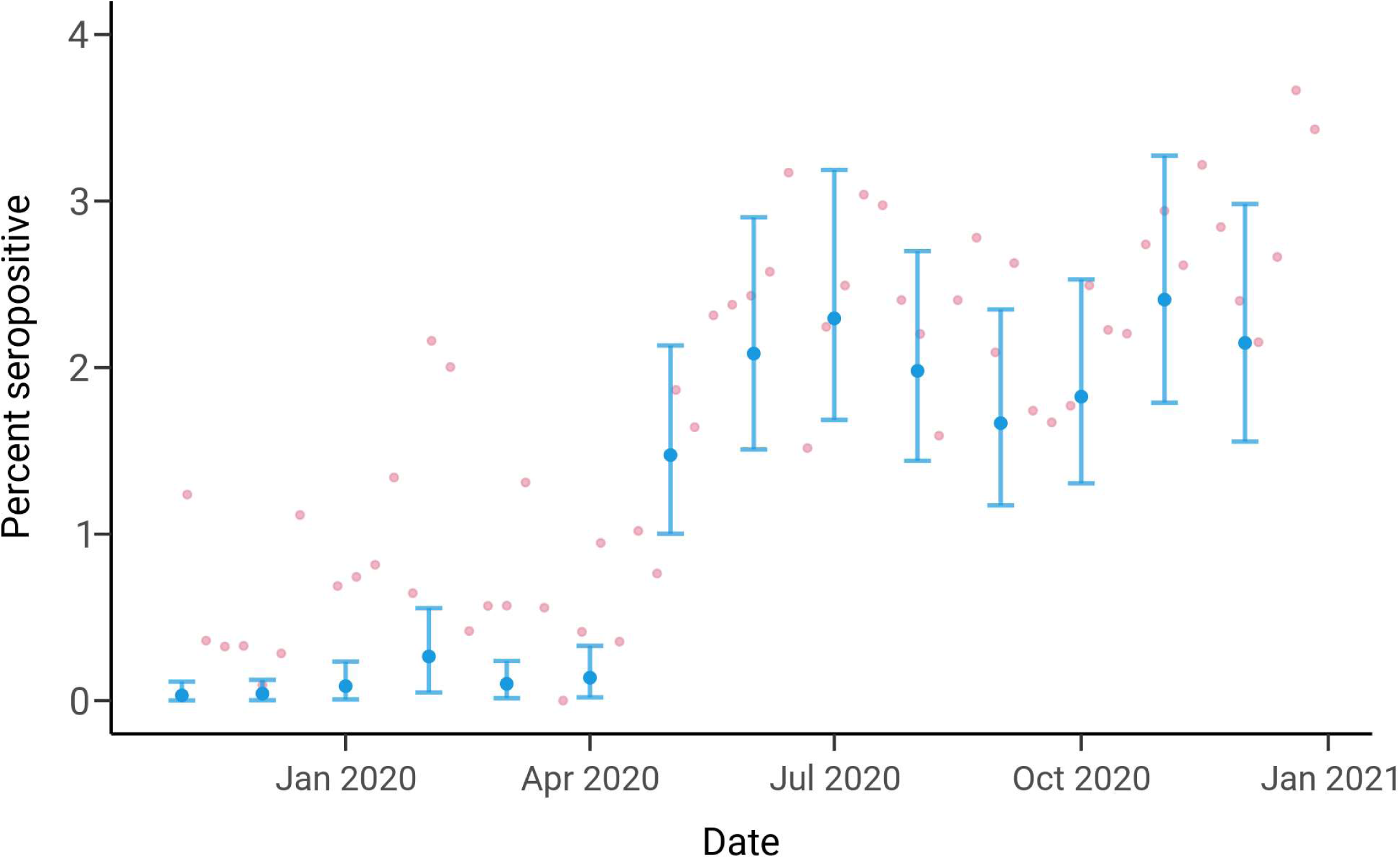
Statewide longitudinal seroprevalence trend from November 2019 to December 2020 estimated using the monthly MRP model adjusting for test specificity. The mean seroprevalence estimates are indicated by the blue dots with the error bars depicting 90% credible intervals; pink dots represent unadjusted weekly seroprevalence estimates.

We next estimated longitudinal seroprevalence trends in Massachusetts cities and towns to identify geographic heterogeneities in SARS-CoV-2 infection risk. Overall seropositivity varied considerably across the state (Figure 3, Supplementary Figure 4), but cities with high seroprevalence showed similar qualitative trajectories to each other and to overall statewide trends (i.e., increases from April to May 2020 followed by plateaus) (Figure 3). The widths of the credible intervals also varied, reflecting uncertainty due to smaller sample sizes for some cities, such as Chelsea, compared to larger cities, such as Boston. The estimate for seroprevalence in Boston was 0.05% (90% CI [0.00, 0.17]) in November 2019 and rose to 3.56% (90% CI [2.49, 4.93]) at the end of 2020, which is slightly higher than the estimate for the state. Modeling time continuously yielded similar qualitative trends (Supplementary Figure 5).

**Figure 3.**
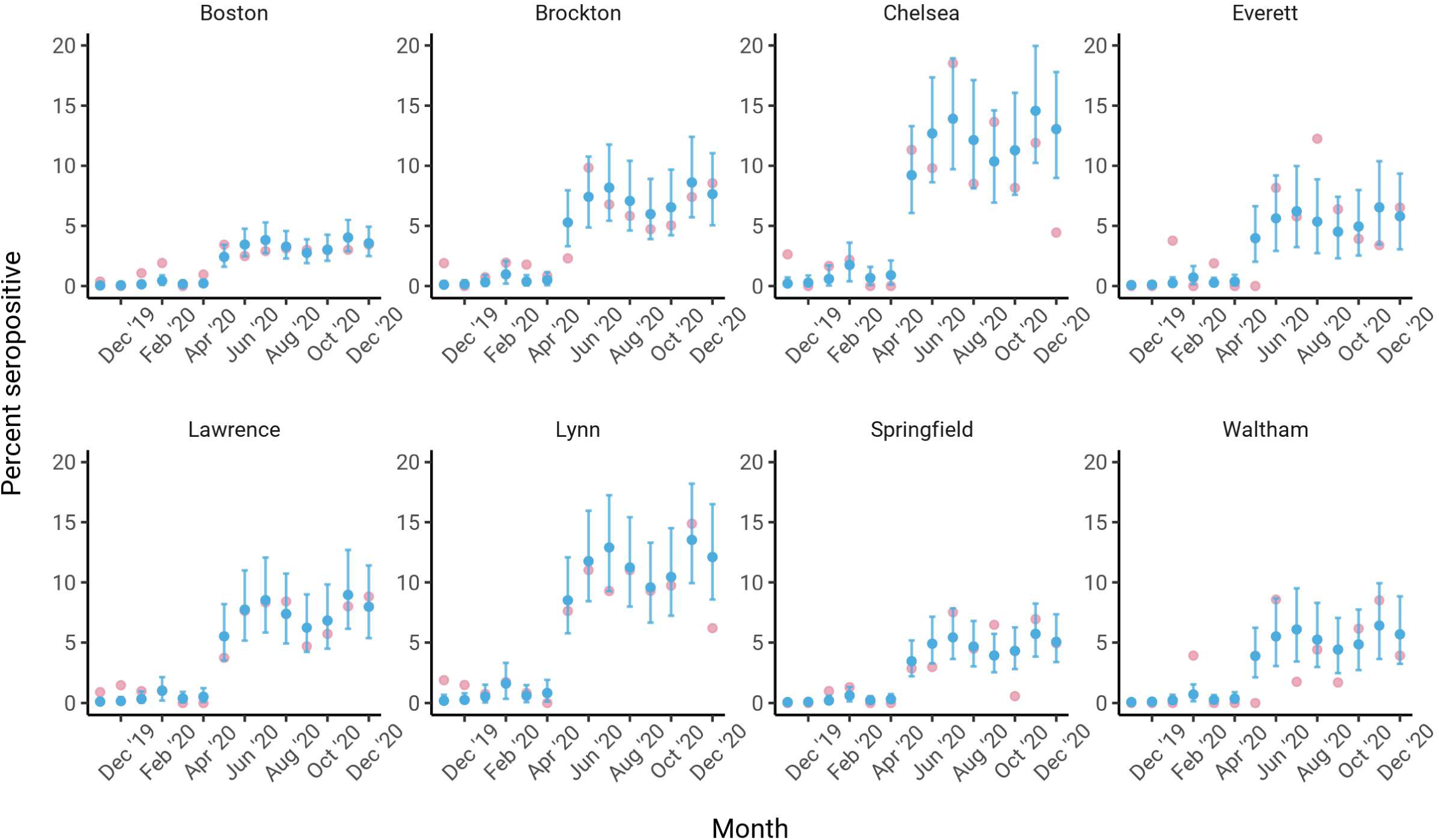
Longitudinal seroprevalence trends in cities and towns from November 2019 to December 2020 estimated using the monthly MRP model adjusting for test specificity. The eight cities and towns with the highest lower 90% credible interval in December 2020 and at least 20 heel stick samples collected are shown. The mean seroprevalence estimates are indicated by the blue dots with the error bars depicting the 90% credible intervals; pink dots represent unadjusted monthly seroprevalence estimates.

We compared the seroprevalence estimates to active case surveillance data collected by the Massachusetts Department of Public Health and wastewater surveillance conducted by the Massachusetts Water Resource Authority (MWRA) and Biobot [13, 14]. Relative to these data – which capture infected individuals shedding virus – seroprevalence levels are a lagging indicator of infection trends because of the delay from infection to antibody production. To account for this, we overlaid the statewide seroprevalence estimates with the MAVEN and MWRA data using a three-week lag (Figure 4). We observed good qualitative concordance between seroprevalence levels and MAVEN cumulative incidence trajectories from 2019 into 2020 but a divergence between the curves at the end of 2020: seropositivity in the heel stick cohort did not rise as sharply as cases did during the winter resurgence in transmission (Figure 4a). This discrepancy held true for trends in selected cities and towns (Figure 4b), as well as comparing seroprevalence levels with cumulative SARS-CoV-2 RNA copies per mL from the wastewater surveillance data (Figure 4c), indicating a deviation between the DBS data and multiple other sources of SARS-CoV-2 surveillance towards the end of the sampling timeline.

**Figure 4.**
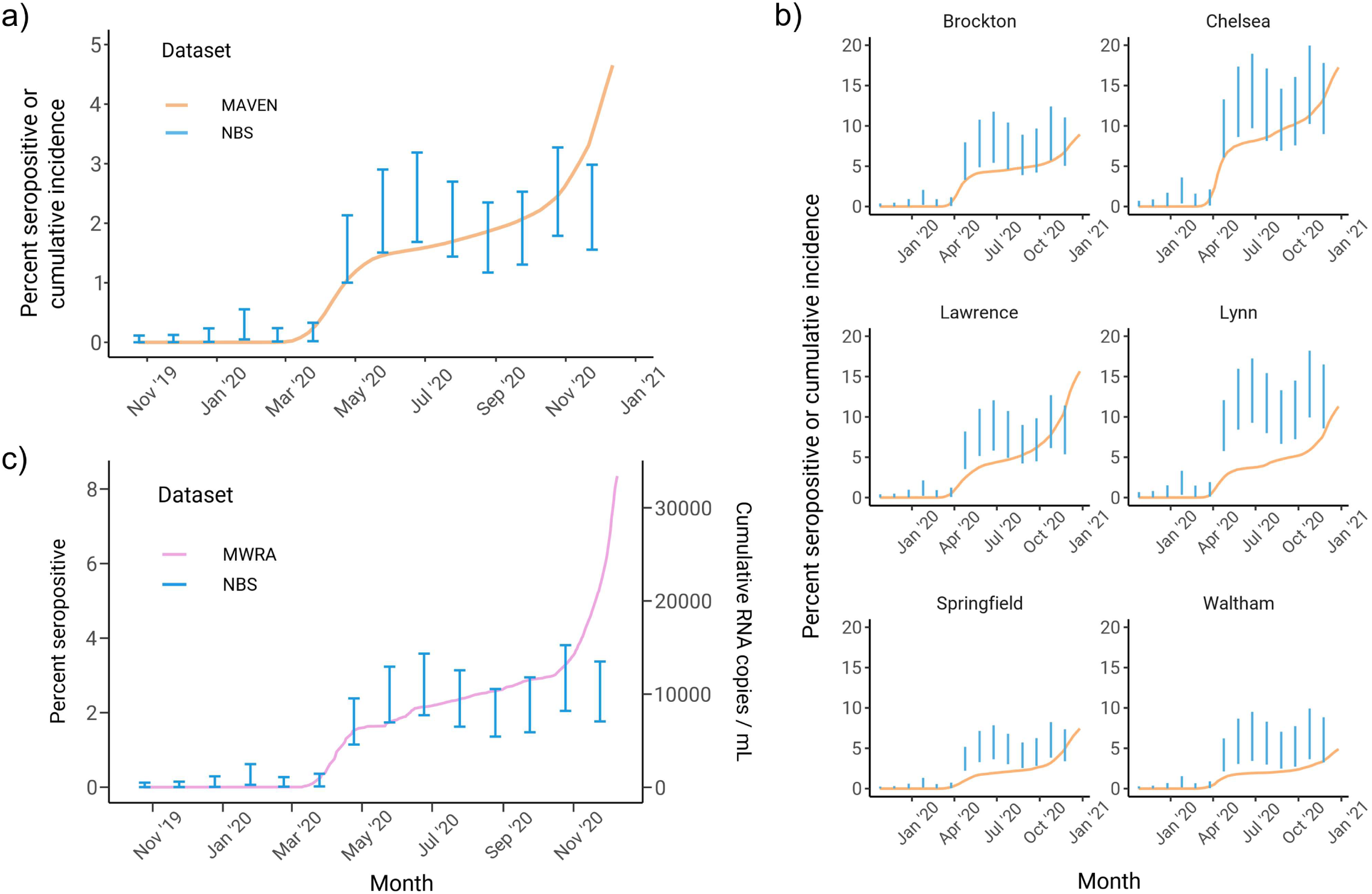
Comparison of MRP seroprevalence estimates with SARS-CoV-2 surveillance data from reported cases and wastewater testing. a) Statewide longitudinal seroprevalence trend estimated from newborn screening samples (blue) overlaid with cumulative incidence from MAVEN epidemiological surveillance data (orange). b) Seroprevalence trends for six cities and towns (blue) overlaid with MAVEN cumulative incidence (orange). c) Seroprevalence trend for the northern MWRA region (blue) versus cumulative RNA copies per mL from wastewater SARS-CoV-2 testing for the same region (pink). In all subpanels, the blue error bars depict the 90% credible interval for seroprevalence, and seroprevalence data are shifted backwards by three weeks to match the timing of surveillance data.

**Figure 5.**
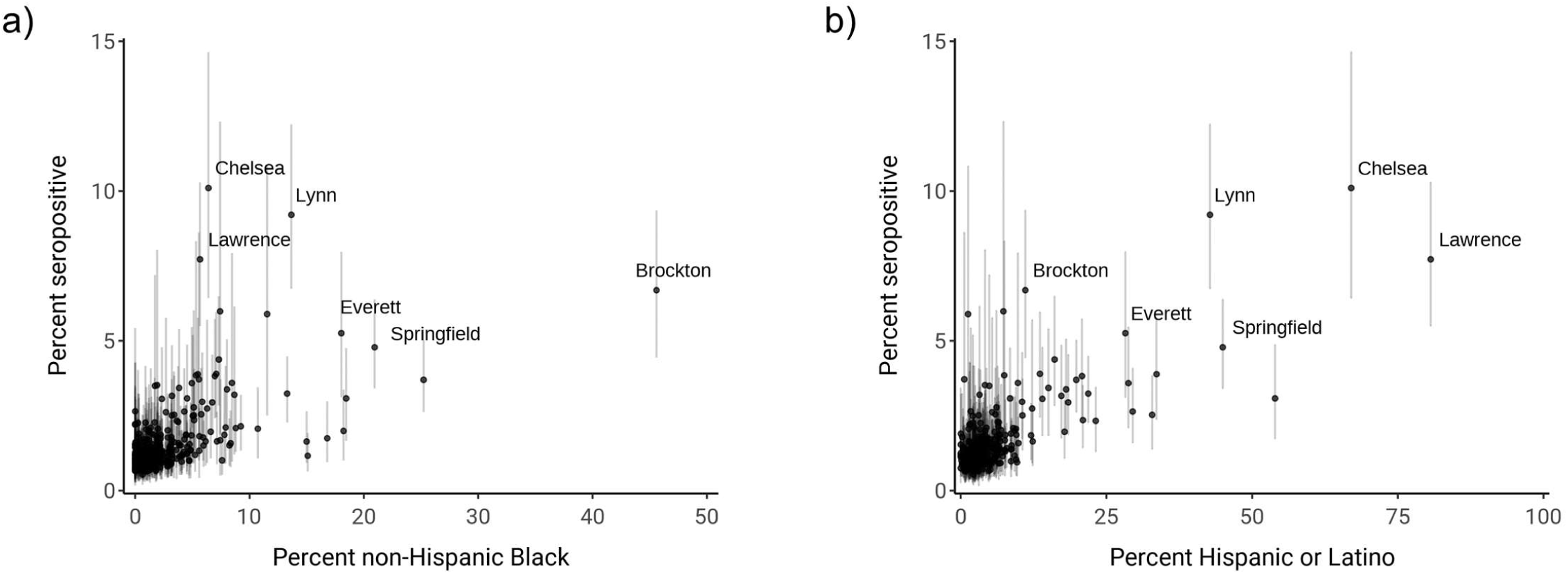
Comparison of percent non-Hispanic Black (left) and percent Hispanic or Latino (right) with estimated seroprevalence by community. Dots indicate mean of the posterior seroprevalence distribution and shaded regions indicate 90% credible intervals. Communities with a lower 90% credible interval above 3% seropositivity are labelled.

Finally, we sought to understand community-level factors associated with increased seropositivity by fitting a multivariate model that included 14 sociodemographic variables on serosurvey data from the last four months. These variables spanned population size and density, race and ethnicity, housing, education, and socioeconomic status (Supplementary Table 2). We calculated 90% credible intervals – representing the most plausible range of values – for the 14 estimated coefficients of association between each variable and seropositivity. The percent of the community that was non-Hispanic Black was the only variable with a 90% credible interval that excluded 0 (Table 1); the mean of the posterior distribution for this variable was 0.024, indicating that a 10% increase in the percent of a community that was non-Hispanic Black was associated with 27% higher odds of seropositivity, adjusting for all other included variables. The association of percent non-Hispanic Black and percent Hispanic or Latino with increased seropositivity was driven in part by several cities and towns, including Brockton, Springfield, Everett, Chelsea, Lynn, and Lawrence (Figure 4). We also calculated the probability of a coefficient being greater or less than zero for the 14 coefficients: percent non-Hispanic Black and percent without insurance coverage had a 90% or higher probability of being greater than 0, whereas average household size, median age, and population density had a 90% or higher probability of being less than 0. These results suggest several additional sociodemographic variables that warrant further study to characterize their association with seropositivity (Supplementary Figure 6).

## Discussion

Understanding the cumulative incidence of SARS-CoV-2 is essential for setting public health policies and guiding vaccine rollouts, but the cumulative incidence for states and their cities and towns is still largely unknown because of a lack of representative serosurveys and inadequate case testing. In Massachusetts, we leveraged the knowledge base and repository of our NBS program for the generation of a readily accessible, retrospective, and de-identifiable source of maternal antibodies that can inform SARS-CoV-2 seroprevalence estimates. The strengths of this approach are multifaceted: the study cohort is free of biases arising from test-seeking behaviors, test availability, and symptom presence, since child-bearing women were included in the study because they had given birth, not because they were thought to have been exposed to SARS-CoV-2 [18]. Additionally, the specimens and demographic data are routinely stored and retrospectively available for public health purposes. The key idea of our study is that by using DBS as the primary data source, we can estimate longitudinal trends in seroprevalence in childbearing women and then project these results to the population at large using statistical methods.

We analyzed data from 72,117 DBS collected across Massachusetts from November 2019 through 2020 and estimated seroprevalence using multilevel regression and poststratification (MRP), which is a Bayesian statistical method used for generalizing survey results and has been increasingly applied to epidemiological studies of SARS-CoV-2 [6, 8, 9, 19]. We found that longitudinal trends statewide and for selected cities and towns were qualitatively similar, with a rise in seropositivity in April 2020 followed by a plateau. The estimated seroprevalence levels had good concordance with cumulative incidence estimated by MAVEN case [13] and MWRA wastewater surveillance data [14], until the observed resurgence of transmission toward the end of 2020. While sampling into 2021 will be necessary to fully understand the extent to which these data sources diverge, one key contributing factor could be changing risk profiles and behaviors of pregnant women over the course of the pandemic. We also found evidence that out of all the sociodemographic variables we investigated, community percent non-Hispanic Black was most associated with increased seroprevalence levels, in line with other epidemiological studies [20-23]. These results underscore the importance of continuing equity and outreach initiatives for minority communities that were most affected by the initial epidemic wave of SARS-CoV-2.

Our findings are subject to at least several limitations and biases. As noted, selection bias could arise because fundamental risk differences between pregnant women and the general population are not accounted for in our statistical model. The direction and causes of this potential bias vary: pregnant women could have less exposure to SARS-CoV-2 due to behavioral choices, depending on sociodemographic characteristics, but increased biological susceptibility to infection due to immune weakening. Nonetheless, estimates from cohorts with clear selection biases, such as blood donors or healthy volunteers, can still meaningfully inform seroprevalence estimates in the general population [18, 24-26]. Second, misclassification bias can occur due to imperfect test sensitivity and specificity. We have estimated specificity using a pre-pandemic sample of DBS and have incorporated it into the statistical model; however, we do not have an estimate for sensitivity. Accounting for imperfect test sensitivity would be expected to shift the seroprevalence estimates higher and widen the credible intervals (Supplementary Figure 7), due in part to the uncertainty involved in measuring sensitivity itself.

By assessing the distribution of maternal antibodies to SARS-CoV-2 statewide and over time, our study provides a strategy for the systematic evaluation and estimation of population-wide cumulative incidence of SARS-CoV-2. Prospective use of NBS-based cumulative incidence estimates of exposure for ongoing policy development would require the more typical same-day turnaround time of clinically-based NBS programs. Our approach – leveraging an easily stored and often readily available data source – may be most useful for informing cumulative incidence estimates in areas where widespread infection testing is still unavailable or remains heavily biased [18].

## Data Availability

These data contain personal identifying health information and are not available for public use or dissemination. Code used for model fitting is available at the following URL: https://github.com/gradlab/covid19-newborn-seroprevalence

## Acknowledgements

We are grateful to Lisa Cavacini PhD, Mass Biologics, UMass Chan Medical School, for providing the CR3022 Mab and Shen Kuang, PhD, Program in Molecular Medicine, T.H. Chan School of Medicine, UMass Chan Medical School for providing RBD protein. We are additionally grateful to Megan Hatch, Lori Chou, Shauna Onofrey, Melody Rush, Arthur Benjamin, Deborah Britton, and the newborn heel stick specimen processing team for laboratory and analytical support throughout the project.

## Supplementary Information

### Supplementary Methods

#### Antibody testing optimization (continued)

Both 96-well (96w) and 384-well (384w) microtiter plate formats were utilized. 3.2mm (d) punches of DBSs were placed into 96-well polyproplylene v-bottom microtiter plates, eluted with 150 µL dilution buffer (1% BSA, 0.05% Tween-20, 140 mM NaCl, 50 mM Tris (pH 8.0)), and incubated ON at 4°C. Final DBS eluates were prepared by transferring the entire eluate from the DBS-containing wells, into clean v-bottom polypropylene microtiter plates. These plates were sealed using Linbro Acetate Sealer from MP Biomedicals,CAT #77-400-05, transported to one of three testing sites, kept at 4°C, and used within 3 days. Flat-bottom polystyrene ELISA plates (Thermo #439454 for 96w; #464718 for 384w) were coated with 100 µl (50 µl for 384w) SARS-CoV-2-RBD (1 µg/ml in CBB buffer; Millipore Sigma (C3041100CAP)) for 30 minutes at RT, and washed X3 in wash buffer (% Tween-20, 400 mM NaCl, 50 mM Tris (pH 8.0)). Wells were then blocked with 250 µl (100 uL for 384w) blocking buffer (1% BSA, 140 mM NaCl, 50 mM Tris (pH 8.0)), for 30 minutes at RT, and washed X3. DBS eluates were diluted 1:4 in dilution buffer, and 100µl (50µL for 384w) was added to wells of the washed, coated, and blocked plates, incubated 30 minutes at 37°C, and washed X5. To each well 100 µL (50 µL for 384w) HRP-conjugated human IgG-specific detection antibody (Bethyl Laboratory (A80-104P)) diluted 1:25,000 in dilution buffer was added. Plates were incubated 30 minutes at RT and washed X5. 100 µL (50 µL for 384w) substrate (1-Step™ Ultra TMB-ELISA Substrate Solution; Thermo Fisher (34029)) was added to each well and incubated 3 minutes at RT, then stopped with 100 µL (50 µL for 384w) 1M H2SO4. Plates were read at 450nm (and background at 570nm subtracted). On each plate, a standard curve of 2-fold serial dilutions of purified CR3022 antibody was performed in duplicate, as well as eluates of DBSs made from blood of adults known to be positive for low concentration of COVID antibody, as controls.

#### Laboratory data harmonization and cutoff determination

To harmonize data among the three testing laboratories, optical densities (OD) of DBS eluates were converted into µg/ml of CR3022 monoclonal IgG antibody by comparing activity to the standard curve. Specifically, a 5-parameter logistic curve function, 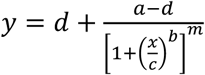, was fit using the curve_fit method of SciPy, a scientific programming package for Python, with x=µg/ml and y=OD [1]. This fit curve was then used to convert OD on a per-sample basis to µg/mL. Converted µg/mL values were then averaged by replicates to obtain a sample µg/mL value. To ensure correct normalization across laboratory and plate, the serial dilution and curve-fitting procedure were performed on every plate.

Between and within laboratories, we observed interplate variability in the reported ODs and concentrations and concentrations attributable to the then single-source low positive control. We investigated a variety of harmonization protocols and determined that a per-plate two cycle cutoff calculation represented data from the three laboratories best. We knew there to be no bias in the distribution of samples from geographic residence locations to each laboratory and, as expected, Supplementary Table 1 shows that the yields of seropositive values from each laboratory are generally proportional to the number of specimens tested by that laboratory.

### Population-wide statistical model (continued)

The formula for the MRP logistic regression model is given by:

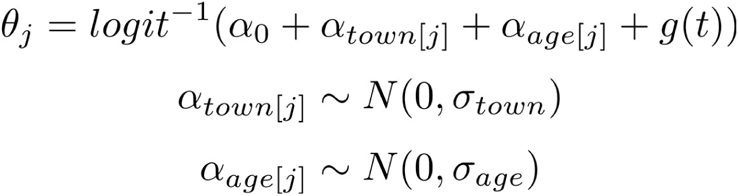

where *θ*_j_ is the seroprevalence in the *j*th strata, *α*_0_ is the intercept, α_*town*[*j*]_ is the city or town random effect for the *j*th strata, *α*_*age*[*j*]_ is the age random effect for the *j*th strata, and *g(t)* is a time-varying function. For the time-varying function, we primarily used estimates from a model grouping data by month:

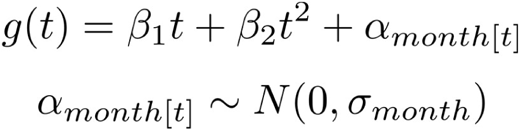

where *t* is the month, β_1_and β_2_ are the coefficients for the overall linear and quadratic month trends respectively, and α_month[*t*]_ is a month random effect for month *t*. We then extend the model with the effect of test characteristics [2]:

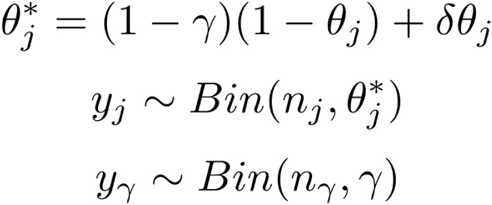

where 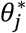 is the test characteristic adjusted seroprevalence for the *j*th strata, γ is the specificity of the test, δ is the sensitivity of the test, *y*_j_ is the observed number of seropositives in the *j*th strata, *n*_j_ is the number of DBS tested in the *jth* strata, *y*_γ_ is the observed number of seronegatives in the pre-pandemic DBS sample, and *n*_γ_ is the total number of DBS tested in the pre-pandemic sample. We assume test sensitivity is fixed at 1 for all analyses, though we have conducted a sensitivity analysis (Supplementary Figure 7) to understand how lowered test sensitivity could affect our results.

We also assessed how different time-varying functions could affect our estimates. To model time continuously, we used a Bayesian generalized additive model with thin-plate splines as in Pouwels et al. [3]. We specified factor smooth interactions, which allow spline parameters to vary by community but with a shared smoothing parameter. This model did not adjust for test characteristics.

### Model fitting and code

The generalized additive model was fit using the stan_gamm function from Rstanarm version 2.19.2 run with 500 iterations, 8 cores, a basis dimension of 5, the smoothing basis set to factor smooth interactions, and otherwise default settings, including priors. The monthly model was fit using custom Stan code run via Rstan version 2.21.2, sampling for 1000 iterations using 4 chains on 4 cores and otherwise default settings. Model fit was assessed using trace plots and the Rhat metric. Posterior distribution estimates were summarized using mean and 90% credible intervals.

---

**Supplementary Table 1.** Proportions of specimens tested and the distribution of positive results from the three laboratories.

|  | Positives | Tested | Pos. per lab of<br>that lab's tests | Tested per lab<br>of total tested | Positives per lab of<br>total positives |
| --- | --- | --- | --- | --- | --- |
| Lab A | 760 | 45,132 | 1.68% | 62.58% | 60.26% |
| Lab B | 299 | 16,714 | 1.79% | 23.17% | 23.71% |
| Lab C | 202 | 10,271 | 1.96% | 14.24% | 16.02% |
| **Total** | **1,261** | **72,117** | **1.75%** | **100.00%** | **100.00%** |

**Supplementary Table 2.**
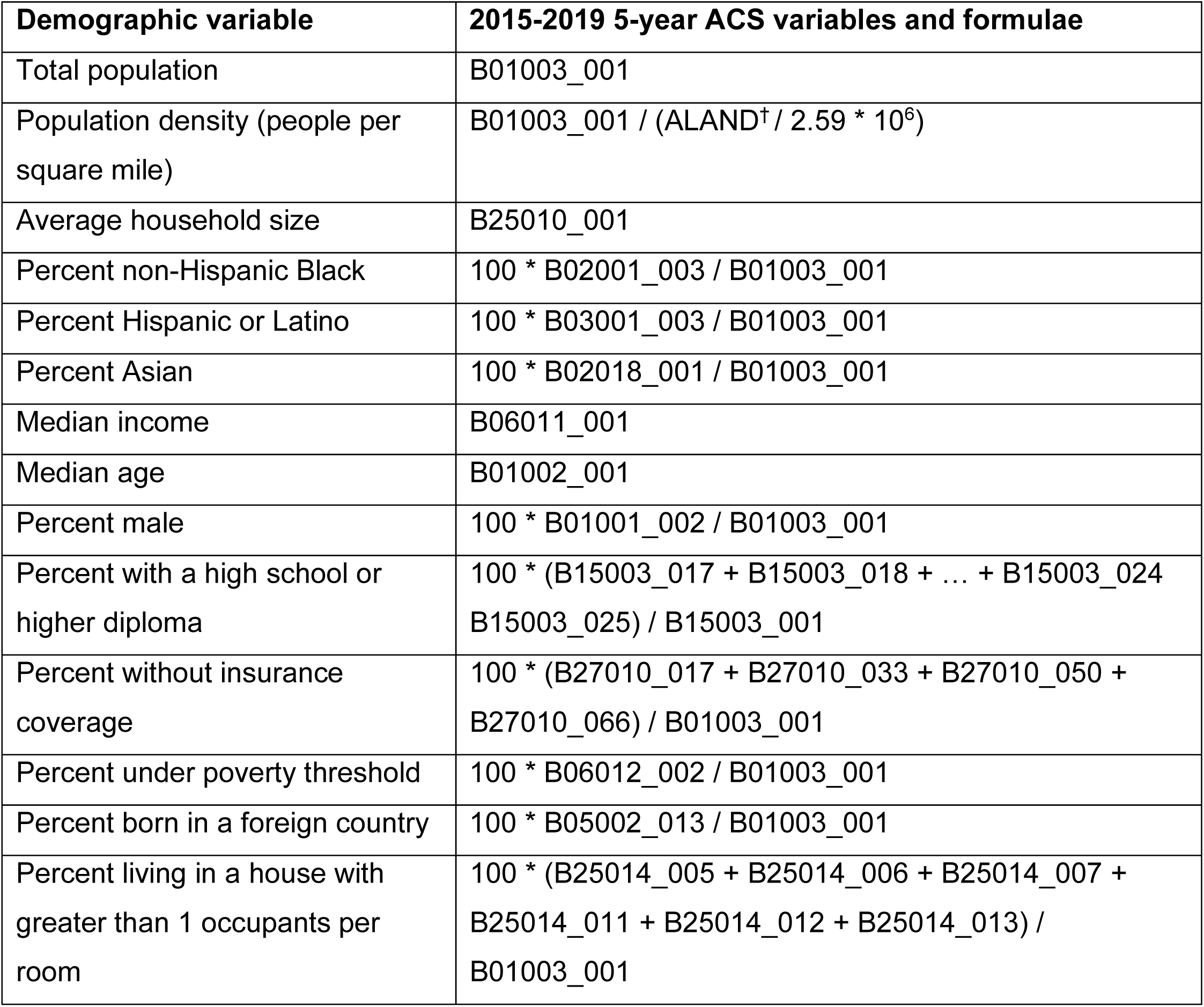
Variables included in the community-level Bayesian multivariate association of demographic characteristics with seroprevalence. †ALAND is the land area in meters squared for each city or town acquired from the US Census Bureau 2018 Cartographic Boundary File at the level of county subdivisions.

**Supplementary Figure 1.**
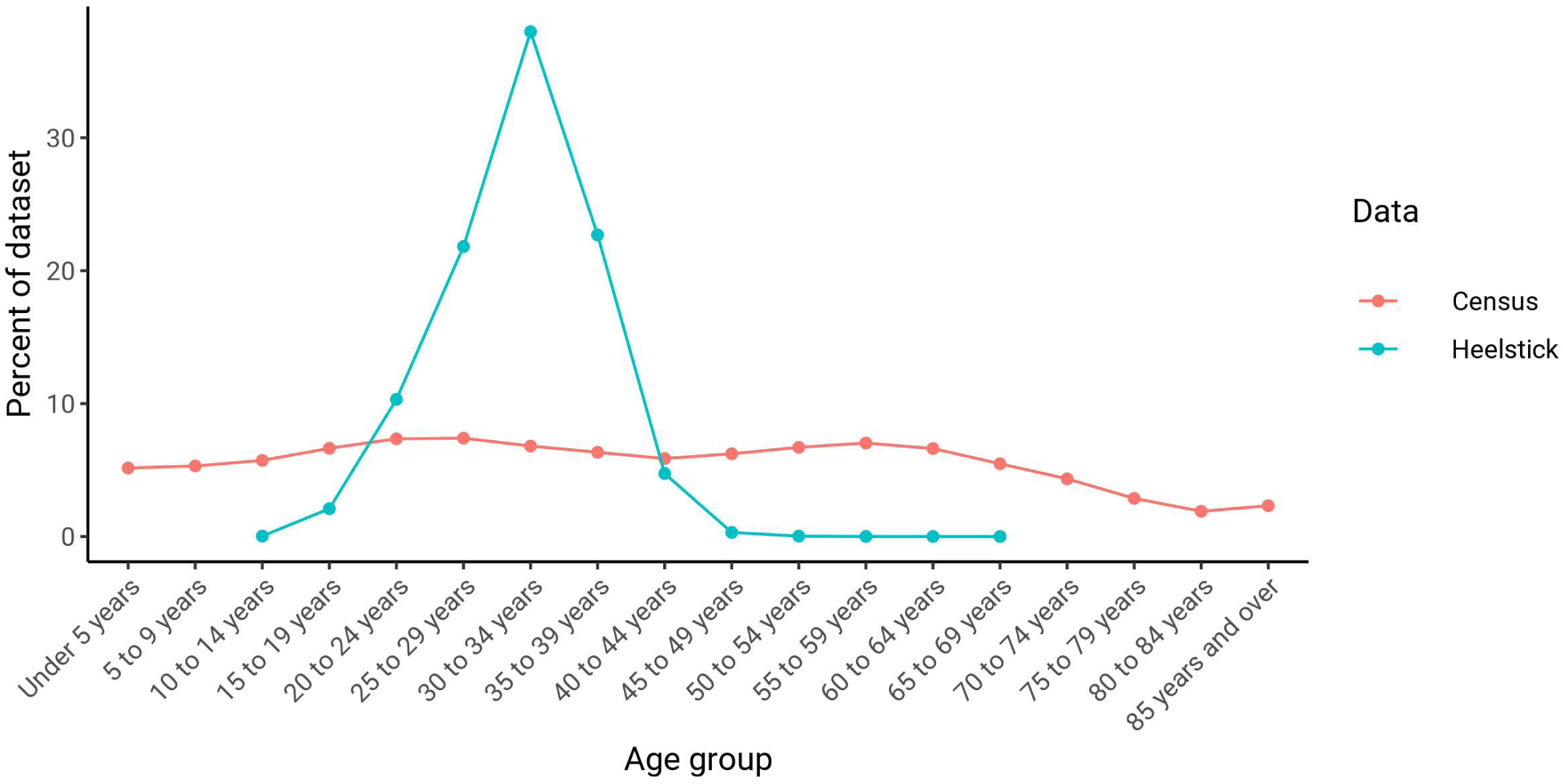
Sampling biases by age in the newborn heel stick data set (blue) compared to census population estimates (red).

**Supplementary Figure 2.**
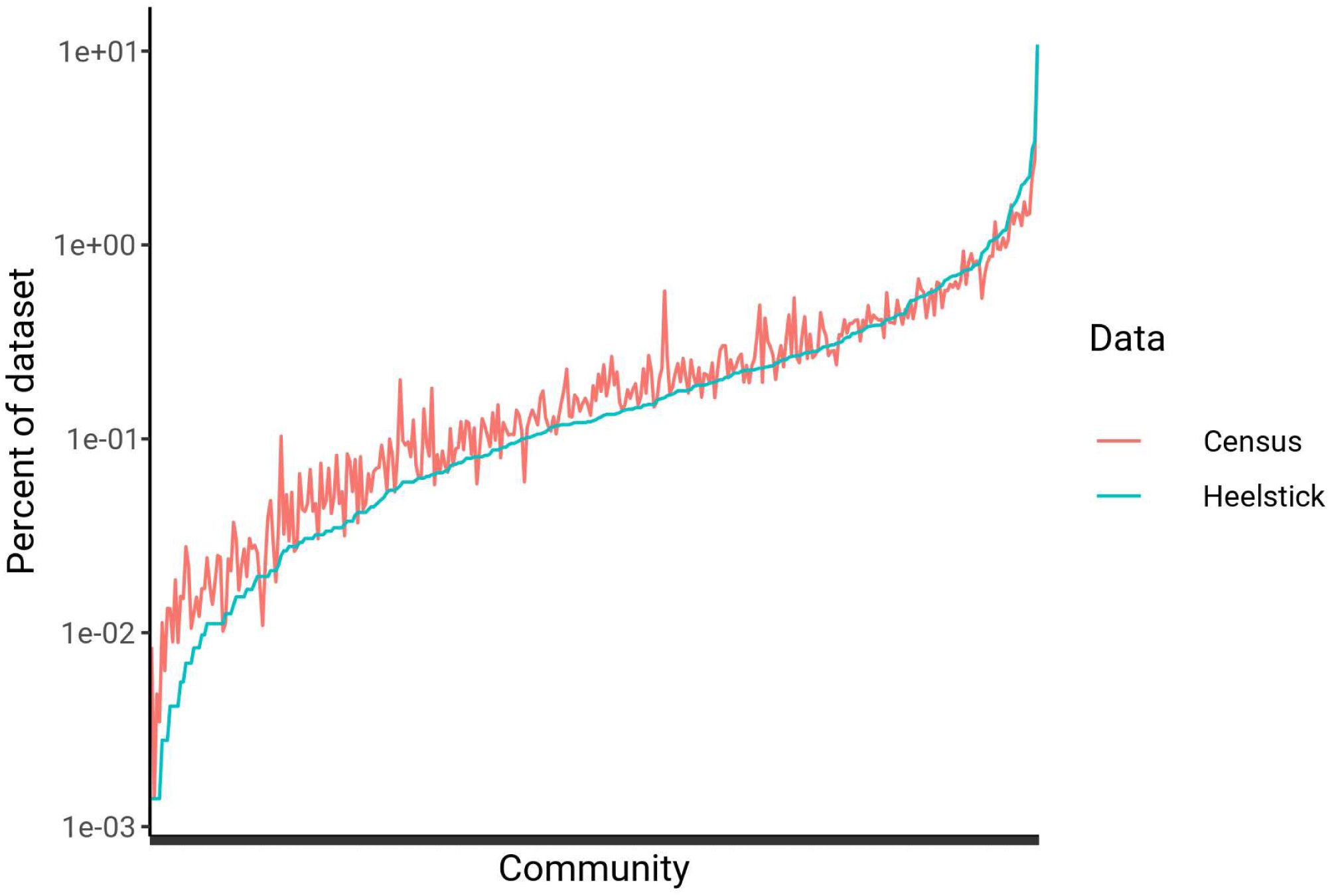
Sampling biases by community (i.e. Massachusetts cities or towns) in the newborn heel stick data set (blue) compared to census population estimates (red).

**Supplementary Figure 3.**
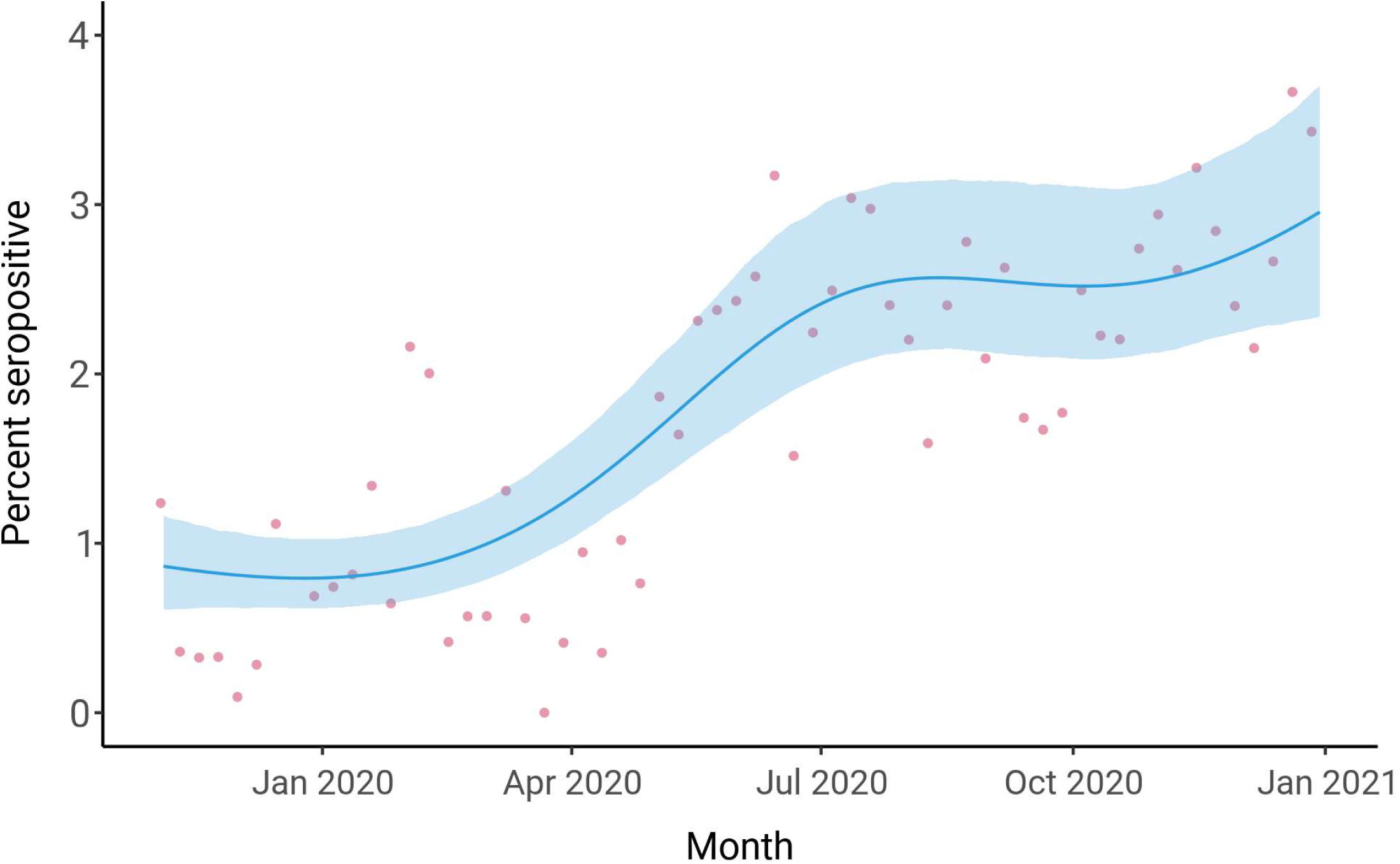
Statewide longitudinal seroprevalence trend from November 2019 to December 2020 estimated using the continuous-time MRP model; test specificity is not adjusted for in this model. The mean seroprevalence estimate is indicated by the blue line with the surrounding shaded region depicting the 90% credible interval; pink dots represent unadjusted weekly seroprevalence estimates.

**Supplementary Figure 4.**
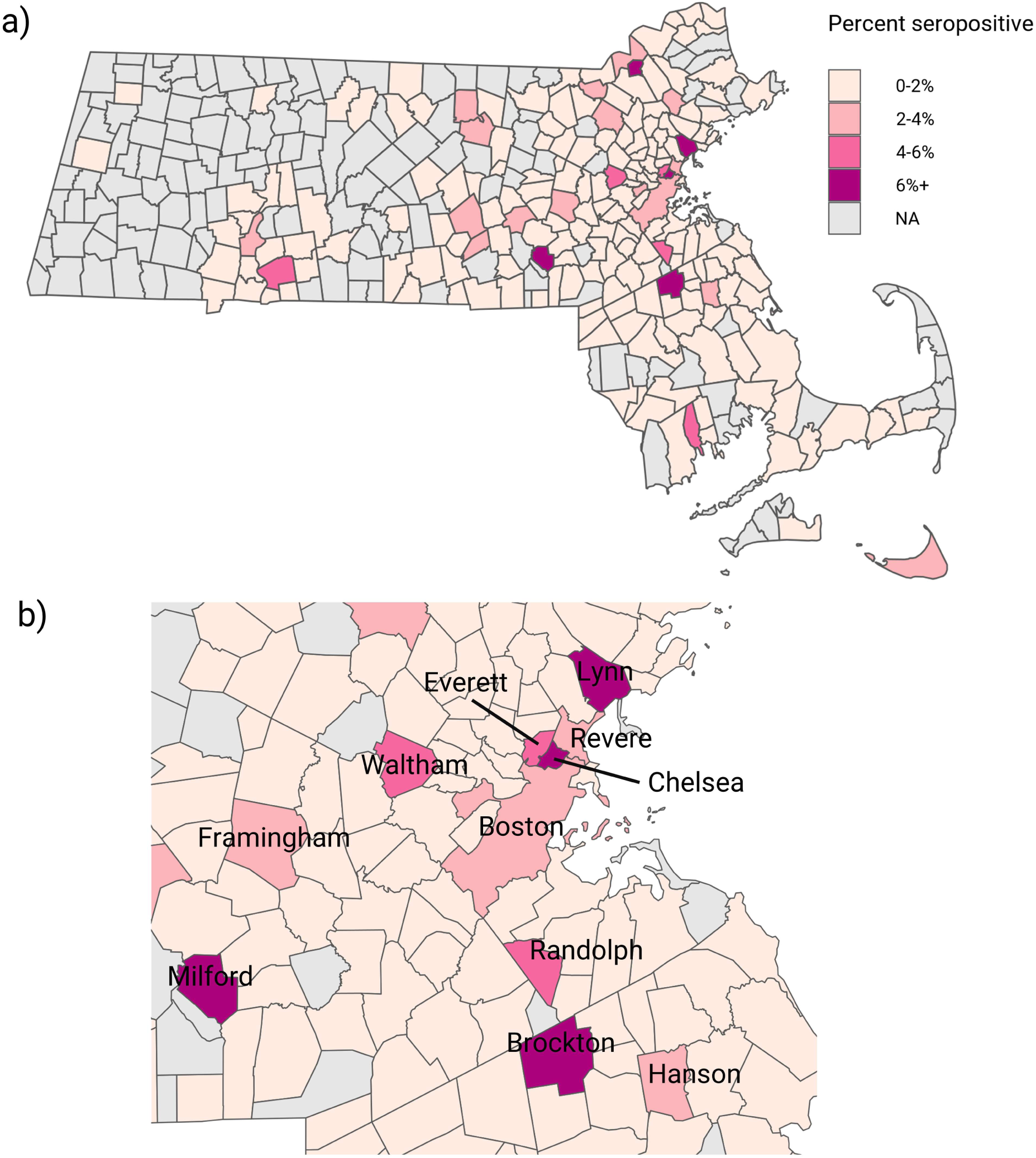
Geographic distribution of seroprevalence by Massachusetts cities and towns statewide (a) and in the Greater Boston area (b). Mean seroprevalence estimated using the monthly MRP model is plotted for December 2020. Cities or towns with less than 5 heel stick samples collected in December 2020 are excluded (NA values).

**Supplementary Figure 5.**
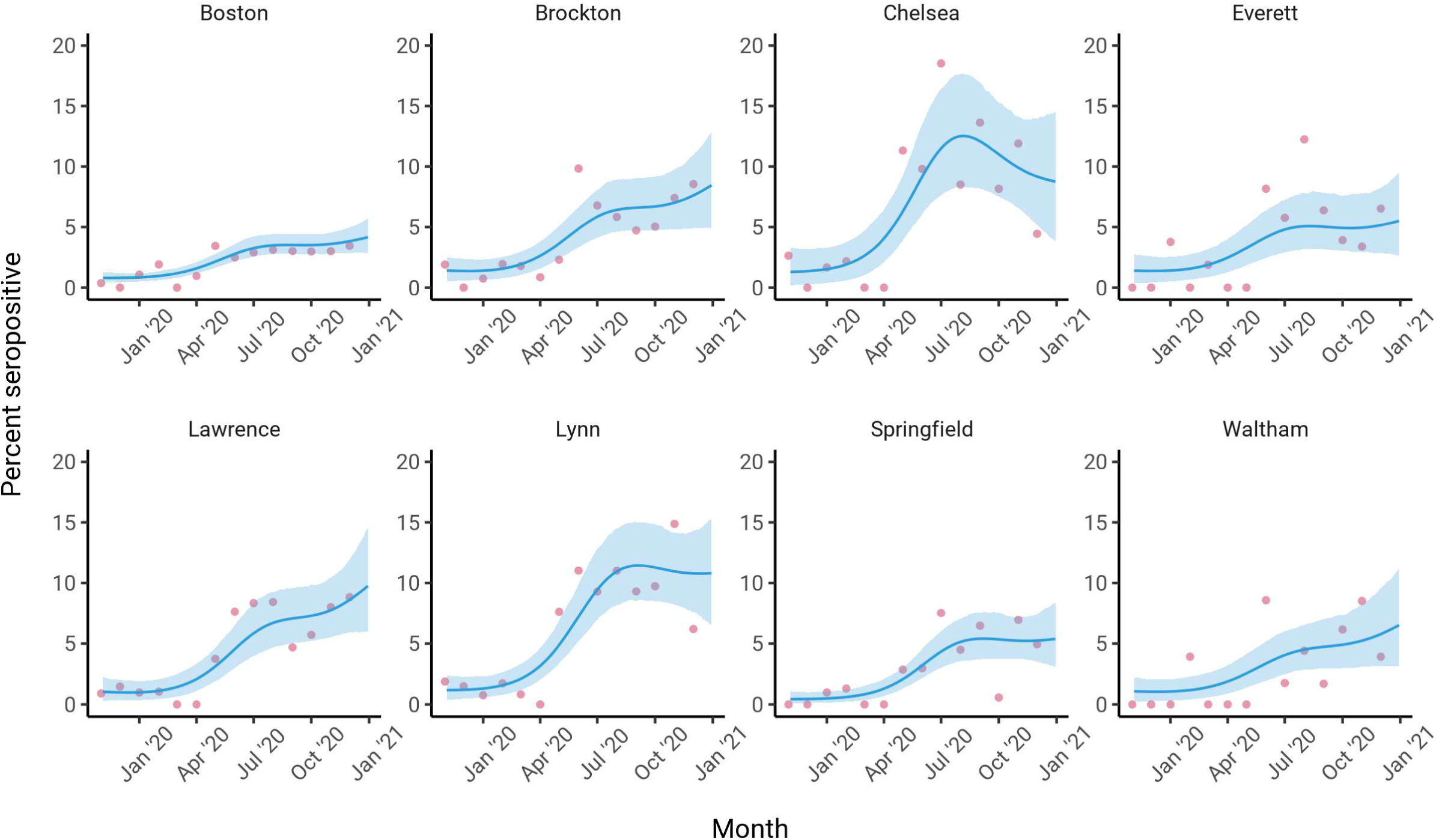
Longitudinal seroprevalence trends estimated through the continuous-time MRP model in the same cities and towns as in Figure 2; test specificity is not adjusted for in this model. The mean seroprevalence estimate is indicated by the blue line with the surrounding shaded region depicting the 90% credible interval; pink dots represent unadjusted monthly seroprevalence estimates.

**Supplementary Figure 6.**
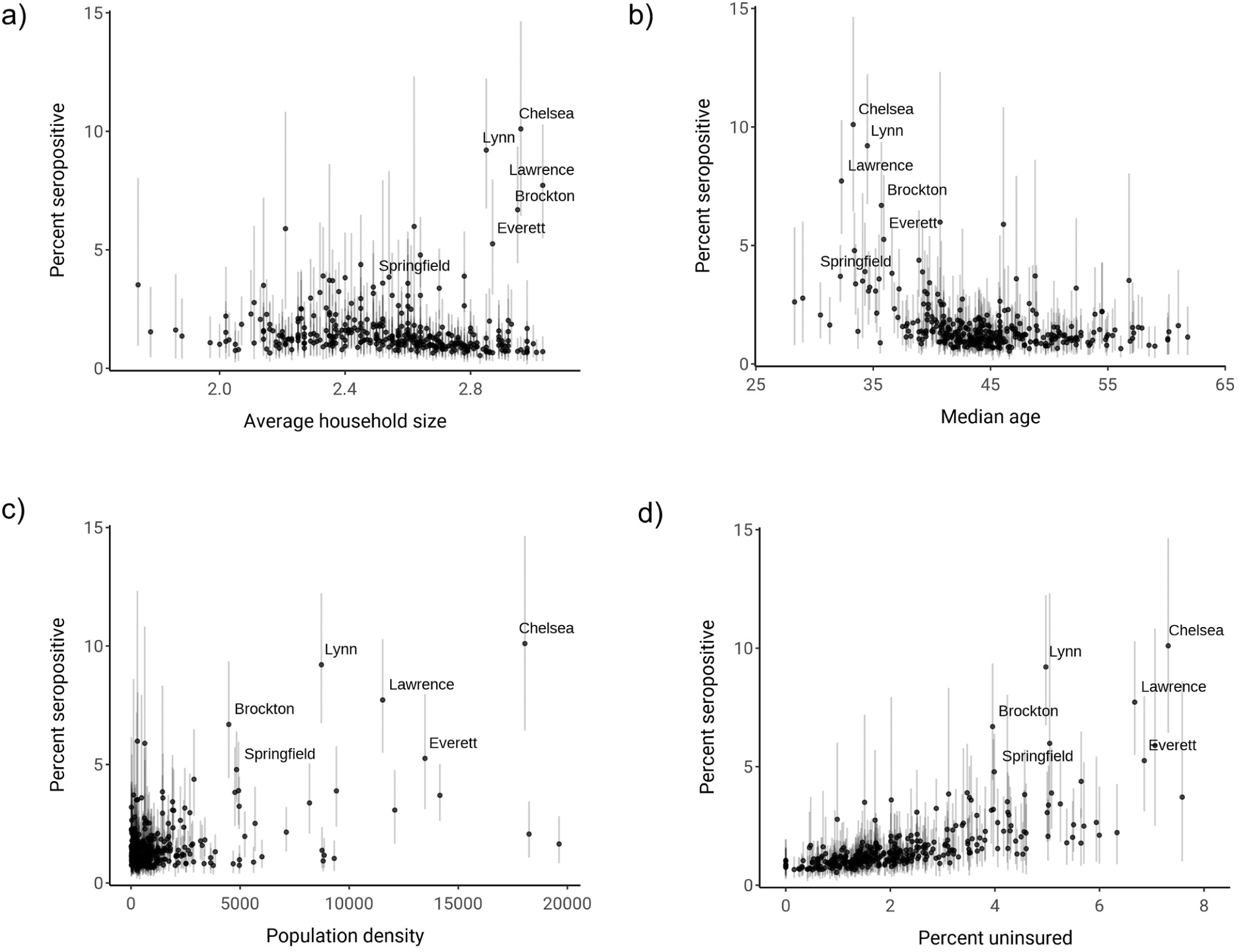
Comparison of a) average household size, b) median age, c) population density, and d) percent uninsured with estimated seroprevalence by community. Dots indicate the mean of the posterior seroprevalence distribution and shaded regions indicate 90% credible intervals. Communities with a lower 90% credible interval above 3% seropositivity are labelled.

**Supplementary Figure 7.**
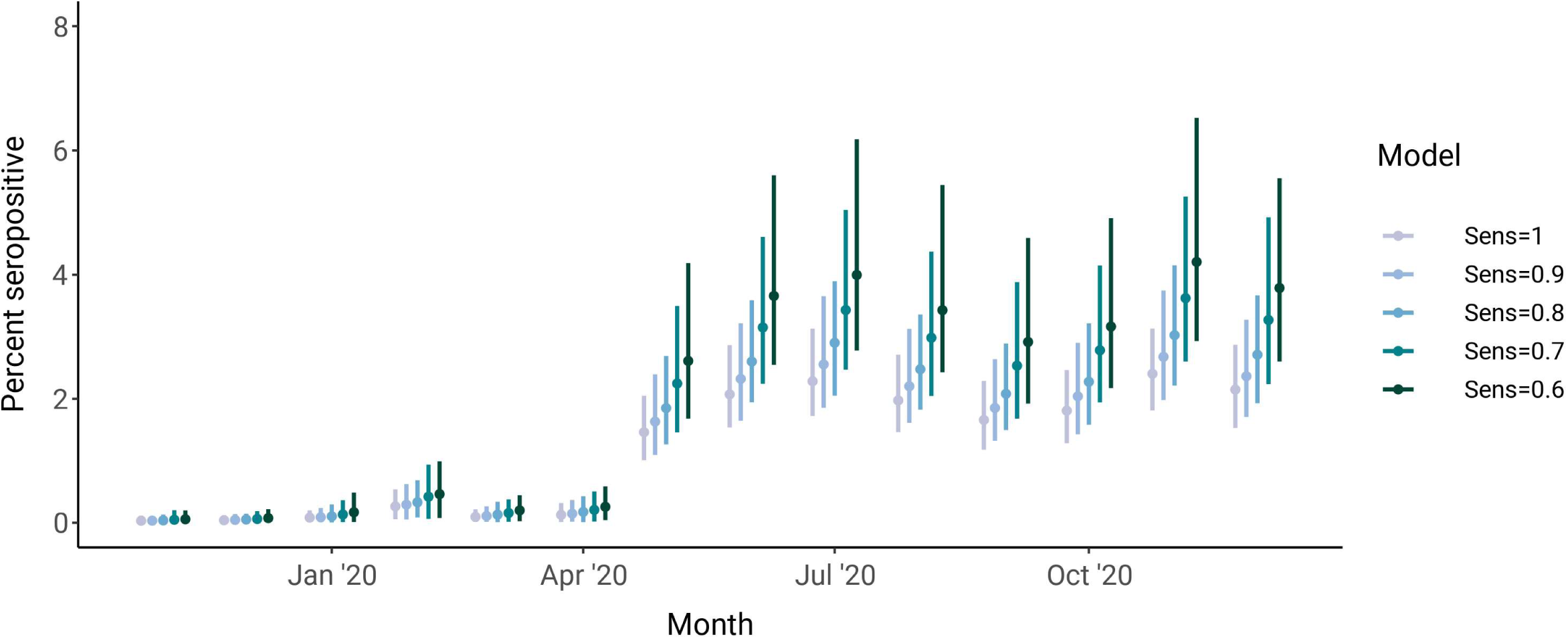
Lowering assumed test sensitivity shifts statewide seroprevalence estimates higher and widens the associated credible intervals.

